# Functional disability, malnutrition and psychological well-being among older individuals: A cross-sectional study in Lalitpur, Nepal

**DOI:** 10.64898/2026.11.11.26367807

**Authors:** Eebaraj Simkhada, Aerusha Simkhada

## Abstract

**Background:** Functional disability, malnutrition, and psychological well-being in older people are deeply interconnected. A better understanding of these conditions is crucial for geriatric care. This study aims to assess the status of functional disability, malnutrition status, and psychological well-being among older people.

**Methods:** Participants were assessed for disability in activities of daily living (ADL) and instrumental activities of daily living (IADL), malnutrition by Mini Nutritional Assessment (MNA) tool, and psychological well-being was evaluated by the WHO-5 Well-Being Index. A multistage cluster sampling probability proportionate to size method was used for sampling, and 360 people aged 60 years and older were selected from the Lalitpur district of Nepal. Data on socio-demographic factors, functional ability, malnutrition, and psychological well-being were collected. Statistical analysis was performed including bivariate analysis to compare genders.

**Results:** The functional disability rate according to ADL was 34.7%, and with IADL disability, it was 47.8%, with higher in women as compared to men. According to the MNA score, this study found a 13% prevalence of malnutrition and 45% at risk of malnutrition. In the WHO well-being index score, 32.8% had a score of < 13, reflecting poor psychological well-being.

**Conclusion:** Nearly one-third of the participants were dependent on ADL, reflecting a high disability rate. About half of the population had poor nutritional status, and nearly one-third of the study sample was experiencing poor psychological well-being. This study findings demonstrate the importance of managing problems with appropriate screening, rehabilitation, and therapeutic measures for the physical and psychological well-being of older people in the community.

## Introduction

Disability is defined as a restriction or inability to perform an activity in the manner or within the range considered normal for a human being [1]. Decreased functional ability in old age is caused by many pathological conditions and an increased burden of comorbidity. Impaired physical function causes restrictions on activities of daily living. Declining physical health can affect the autonomy of people. Difficulty in activities of daily living can affect family life, social relationships, and psychological health. Likewise, malnutrition in the elderly is a challenging health concern due to its effects on elderly health. It is associated with morbidity and mortality in older people, and it is associated with disability and the physical well-being of the elderly population [2].

### Functional Disability in older individuals

Disability is not only musculoskeletal impairment but is an ecological gap between the environmental demands of a person and that person’s capability. Elderly functional capacity may be reduced by impairments in any body system, including the cardiovascular system, musculoskeletal system, and neurological system [3].

Disability is becoming of increasing importance as populations age, and it is a global public health concern. It affects people of all ages and all socioeconomic backgrounds; however, high rates of disability are seen among elderly people [4].

Disability is a loss of functional ability due to the complex interaction of health, environment, and personal factors. Disability, frailty, and comorbidity are used synonymously in the elderly; however, these three terms are distinct clinical entities that are causally related [5]. Frailty, disability, and multi-morbidity are interrelated conditions [6].

Frailty is a state of high vulnerability for developing increased dependency and mortality when exposed to a stressor for adverse health outcomes, including falls, hospitalization, physical disability, and death [7].

Global prevalence of disability is 16%, affecting people of all ages [8]. Disability rates are higher in the older population globally [9]. Due to the increased global burden of non-communicable diseases, [10] and other age-related comorbidities [11], disability in the elderly has become more complicated. Moreover, people with disabilities are facing the social stigma of disability, loss of role in the family, unemployment, and poor healthcare [12–13].

Prevalence of disability based on the National Demographic Health Survey [14] in Nepal was 27% in 2022. A higher disability rate, about 45%, was seen in people aged 80 years and above in Nepal [4].

Components of disability such as, impairments, activity limitations, and participation restrictions, lead to multiple forms of discrimination, predisposing individuals to injury, abuse, and violence [15].

Reducing disability among the elderly is an important issue for their health and well-being. Impaired physical function causes restrictions on activities of daily living. Difficulty in activities of daily living can affect family life, social relationships, psychological health, and feelings of loneliness [16].

### Malnutrition in older people

Malnutrition is a significant health concern for older adults, affecting their overall well-being and quality of life. Malnutrition in the elderly is driven by appetite loss (anorexia of aging), chronic diseases, social isolation, and chewing/swallowing difficulties [7]. Key signs include unintentional weight loss, loose clothing, low energy, muscle weakness, fatigue, or irritability and frequent illness or infections requiring, in some cases, intervention with nutrient-dense foods or oral supplements [17].

Nepal is one of the countries having a high prevalence of malnutrition [18]. In Nepal, a 19.2% prevalence of underweight among adults was reported in NDHS 2016 [19]. Older adults are more likely to be affected by malnutrition than young persons. The prevalence of malnutrition among older people is even higher in Nepal. It is reported to be about 10 to 30% in various studies (20, 21, 22).

Malnutrition causes ill health, functional dependence, and increased susceptibility to complications associated with chronic illness. In older individuals, variables such as gender (women being at higher risk) and having mental disorders were implicated for malnutrition [23].

### Psychological well-being

The World Health Organization defines psychological well-being as a state of mental well-being that enables people to cope with the stresses of life, realize their abilities, learn well and work well, and contribute to their community [24]. It is characterized by feeling good and functioning effectively, going beyond just the absence of mental illness to include having a sense of purpose, meaning, self-acceptance, personal growth, positive relationships, autonomy, and the ability to manage one’s environment and life challenges [25]. Psychological well-being is a multidimensional construct that reflects the ability to grow, adapt, and maintain positive relationships throughout life, particularly for older adults [26]. It encompasses self-acceptance, autonomy, and effective coping with life situations, with high well-being linked to factors such as social support and lower levels of depression, stress, and anxiety [25]. These factors contribute to individuals’ psychological well-being, contentment, and happiness. A six-factor model of psychological well-being was developed by Carol Ryff from diverse well-being theories and research [27]. This model indicates that a good life is balanced and whole; it is not only based on happiness and positive emotions but should also involve engaging in each of these different aspects of well-being.

Psychological well-being is linked to lower mortality risk and better physical health. It provides greater resilience and helps in coping better with stress and setbacks. It provides higher life satisfaction as well as contributes to overall contentment and fulfillment [28]. Studies have shown that psychological health variables such as positive emotions, life satisfaction, optimism, life purpose, and social support were associated with the good health of an individual [29]. In Nepal, a study reported that resilience, self-efficacy, social support, and social connectedness were the main positive factors associated with psychosocial well-being among older adults [30].

### Rationale of the study

Factors causing adverse health outcomes in older people are age, frailty, disability, institutionalization, malnutrition, and mental health [31]. It is evidenced that geriatric assessments reduce nursing home admission, the risk of falls, and the risk of hip fractures, reduce pressure sores, and decrease the risk of delirium in hospitalized patients [32]. Studies have indicated that comprehensive geriatric assessments (CGA) improved health outcomes in older people. There is a gap in knowledge on these aspects of health of older adults in Nepal with scarce data availability.

Hence, the main components of geriatric assessments, functional disabilities, mental health conditions and malnutrition, if recognized at an early stage in older population, can be managed properly.

Information regarding these conditions in the community is necessary to plan and deliver healthcare services in this area. Therefore, it is of the utmost importance to assess the burden of functional disability, malnutrition, and psychological well-being in the older population of Nepal.

The research question was: what is the prevalence of functional disability, malnutrition, and poor psychological well-being among older people?

The objective of the study was to describe the status of functional disability, malnutrition, and poor psychological well-being and to observe the gender differences among the elderly in Lalitpur, Nepal.

## Methods

### Study design and participants

This community-based cross-sectional study was conducted among persons aged 60 years and above in the urban area of Lalitpur District, Nepal. Multi-stage cluster sampling with probability proportional to size was used to select the study sample. This study used the old political structure of VDCs and municipalities, prior to the restructuring of federal boundaries in the districts of Nepal which took place in 2017 [33].

The calculation of the sample size was carried out taking a 31% prevalence rate of malnutrition among home-living elderly people, based on the results of different studies [22, 34–35]. A sample size of 360 was obtained using Epi Info version 7 (StatCalc) with the formula for a population survey based on the following assumptions: prevalence = 31%, confidence limit = 7%, design effect = 2, and confidence level = 95%.

The following were the inclusion criteria for the study: older people aged 60 years or older, who were residing in the area for at least the last 6 months, and those who consented to participate in the study. This study was conducted between September and November 2014. Sampling methodology has been described in detail in our previous paper [61].

### Data collection and study variables

Training was provided for the enumerators on study methodology, questionnaires, data collection methods, record keeping, and proper documentation. House-to-house visits were conducted in the selected clusters to collect the data. In cases of inability to communicate effectively due to language or speech difficulties, which led to difficulty in understanding, the help of a family member was taken. The personal data of the participants were anonymized for the purpose of data privacy protection.

The interview questionnaire was developed in English, translated into Nepali, and then back-translated into English to check the correctness of the translation. It was administered to participants in Nepali by trained interviewers. Pre-testing of the questionnaire with older individuals was done before administration in the study population, and minor grammatical errors were corrected for clarity.

### Functional disability

Activities of Daily Living assess an individual’s ability to perform basic life activities. This scale includes questions in relation to completing everyday activities such as walking, getting in and out of bed, sitting down on or rising from a chair, bending down to pick something up, taking a bath, combing hair and shaving, dressing/undressing, using the toilet, and eating. Activities of Daily Living is a tool commonly used in multidimensional geriatric assessment developed by Katz [36]. ADL explores disability in six dimensions of daily living: bathing, dressing, using the toilet, movement inside the home, feeding, and continence. Continence was considered more of an impairment rather than a disability. It was not included in the scale because bowel/bladder incontinence reflects an abnormality in a particular physical system [37]. The responses were scored on a 4-point Likert scale ranging from “0” (Cannot do at all) to “3” (No problem). The scoring system gave a score range from 0 to 5. Disability was defined as follows: an ADL score of 0, being independent for all items, for the absence of disability; an ADL score of 1, dependent for one item, for moderate disability; and an ADL score ≥ 2, dependent for two or more items, for severe physical disability, based on previous studies [38, 39].

The IADL included 6 activities: shopping for groceries, difficulty with the preparation of meals, difficulty with laundry work, difficulty with light housework, difficulty with travel by public transport, and difficulty with taking the correct dose of medicine [40]. Two items of the IADL scale, use of the telephone (look up the number, dial, and answer) and management of money (write checks and pay bills), were not included because these are not applicable to the majority of Nepalese elderly [16]. The responses were scored on a 4-point Likert scale ranging from 0 (Cannot do at all) to 3 (No problem). For disability analysis, responses were categorized as: score = 0, being independent for all items, for absence of disability; IADL score 1, dependent for one item, for moderate disability; and IADL score ≥ 2, dependent for two or more items, for severe physical disability.

### Malnutrition

The Mini Nutritional Assessment tool (MNA) was applied to evaluate malnutrition in the elderly population. The study employed the full version of the Mini Nutritional Assessment (MNA). The MNA has been validated with clinical, anthropometric, and biologic variables in the geriatric population with a sensitivity (the ability to detect malnutrition) of 96%, specificity (the ability to detect well-nourished status) of 98%, and a predictive value of 97% [41]. The measurement of the MNA variables in the study is described in detail in our previous paper [61].

### Psychological wellbeing

The World Health Organization (WHO) well-being index, the WHO-5A, was used to assess the psychological well-being of the study population, as used in previous studies and also applied successfully across a wide range of study fields [42, 43]. It is a 5-item scale, and the score is calculated by totaling the figures of the five answers. The score ranges from 0 to 25, with 0 representing the worst possible quality of life and 25 representing the best possible psychological well-being and quality of life. It assesses the subjective well-being and state of mood of the participants. It allows the detection of depression if the score is less than 13. As a sensitive and specific tool, it has adequate validity both as a screening tool for depression and as an outcome measure balancing the wanted and unwanted effects of treatments in clinical trials [44].

### Socioeconomic variables

Sociodemographic characteristics of the study population, such as information on age, gender, ethnicity, place of residence, marital status, economic status, education, and occupation, were recorded.

**Age:** Age in completed years; categorised as: 60 to 69 years, 70 to 79 years, above 80 years

**Sex:** Male and female

**Marital status:** Married, unmarried, widowed, divorced

**Education:** Respondent’s highest level of education categorised as:

Illiterate (unable to read and write)

Informally literate: Literate with informal education Primary: Formal education up to five classes

Secondary and above: Formal education six classes and above

**Ethnicity:** Classified as Brahmin/Chhetri, Newar, other Janajati (Magar, Tamang, Gurung), and Dalit.

**Main current occupation:** Recorded as farmer, laborer, self-employed/job, and housework/no work.

**Economic status:** Poorest, poor, middle, richer, and richest income groups.

Economic status was assessed by the Department of Health Services (DHS) weighted wealth index method [45]. Housing characteristics and household assets data were obtained. Factor analysis was performed using the principal component analysis method, and the household’s wealth index value was calculated. Then, the index was categorized into: lowest as poorest, second as poor, middle as middle, fourth as richer, and highest as richest income groups.

### Data processing and statistical Analysis

Data management was carried out in Microsoft Excel 2007, and the statistical analyses were conducted using the Statistical Package for the Social Sciences (SPSS 20.0). Descriptive analysis was performed to assess the characteristics of all variables. Frequencies and percentages were calculated for categorical variables. The prevalence of functional disability, malnutrition, and psychological well-being was estimated and reported as prevalence with 95% CI (confidence interval). Chi-square and F tests were used to assess differences between genders on variables (socio-demographic factors, health characteristics, functional status, and psychosocial status), and a p-value of < 0.05 was considered statistically significant for all analyses.

### Ethical consideration and consent

The Institutional Review Committee, Institute of Medicine, Maharajgunj Medical Campus, Kathmandu, Nepal, approved this study (Reference no. 75(6-11-E)071/072). Before data collection, written informed consent was obtained from all participants, and thumb impressions were obtained from illiterate participants.

## Results

Among the 360 elderly people selected for the study, the sample included 183 women (50.8%) and 177 men (49.2%). The mean age was 71.2 (SD=8.2) years and was similar in both genders.

Table 1 presents the sociodemographic profile of the study population distributed by gender. Ethnicity-wise, Newars comprised 68% of the participants, forming the major group in the study. More older people were widowed (41.1%), with a higher proportion in the women’s group (61.2%). In educational status, 67.8% were illiterate, and women were more illiterate as compared to males. About 14.4% did not have formal education but were literate, 6.9% had completed primary level education, and 10.8% had secondary level or higher education. In the occupation category, approximately 71.7% were farmers by occupation in the study sample.

**Table 1.**
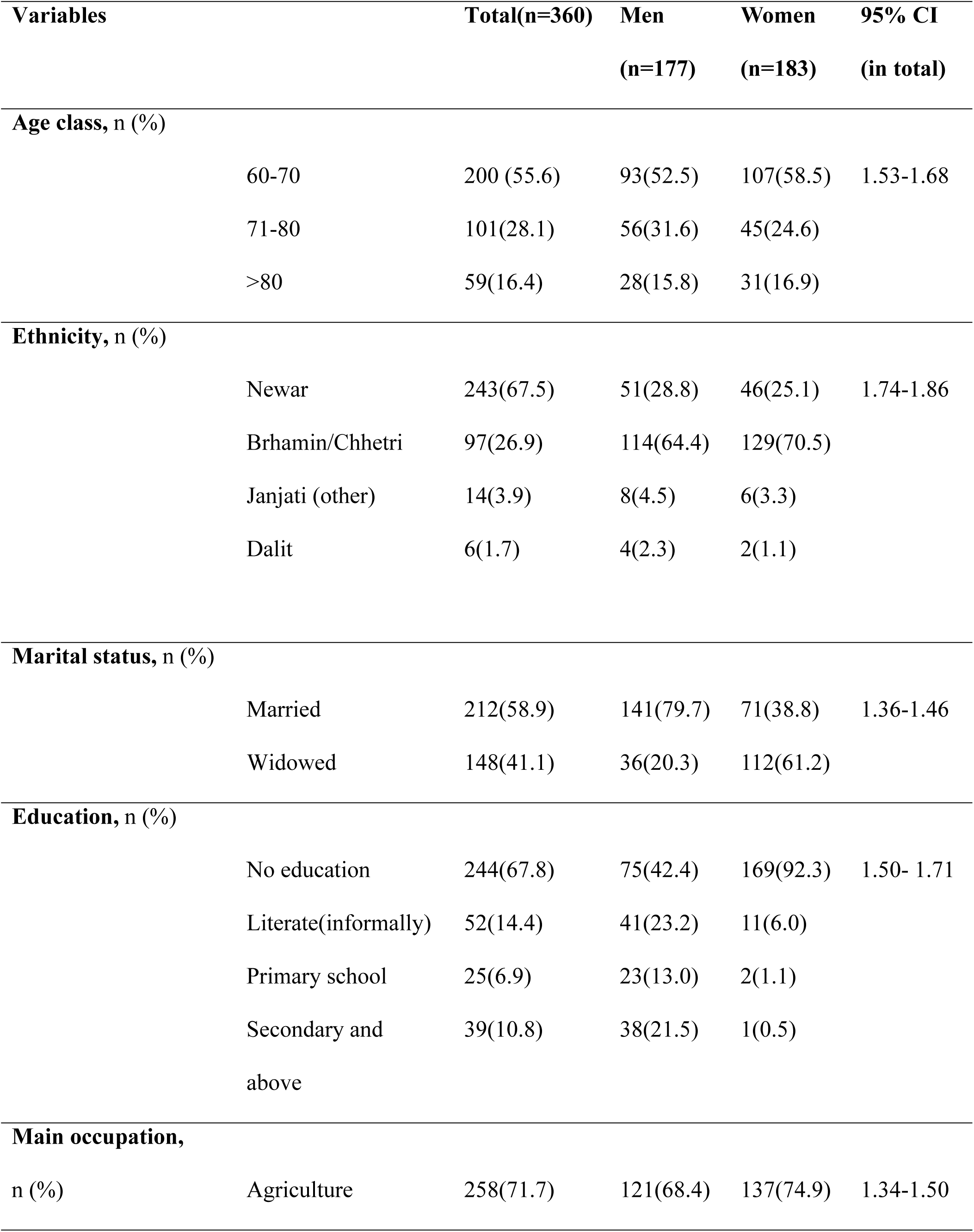
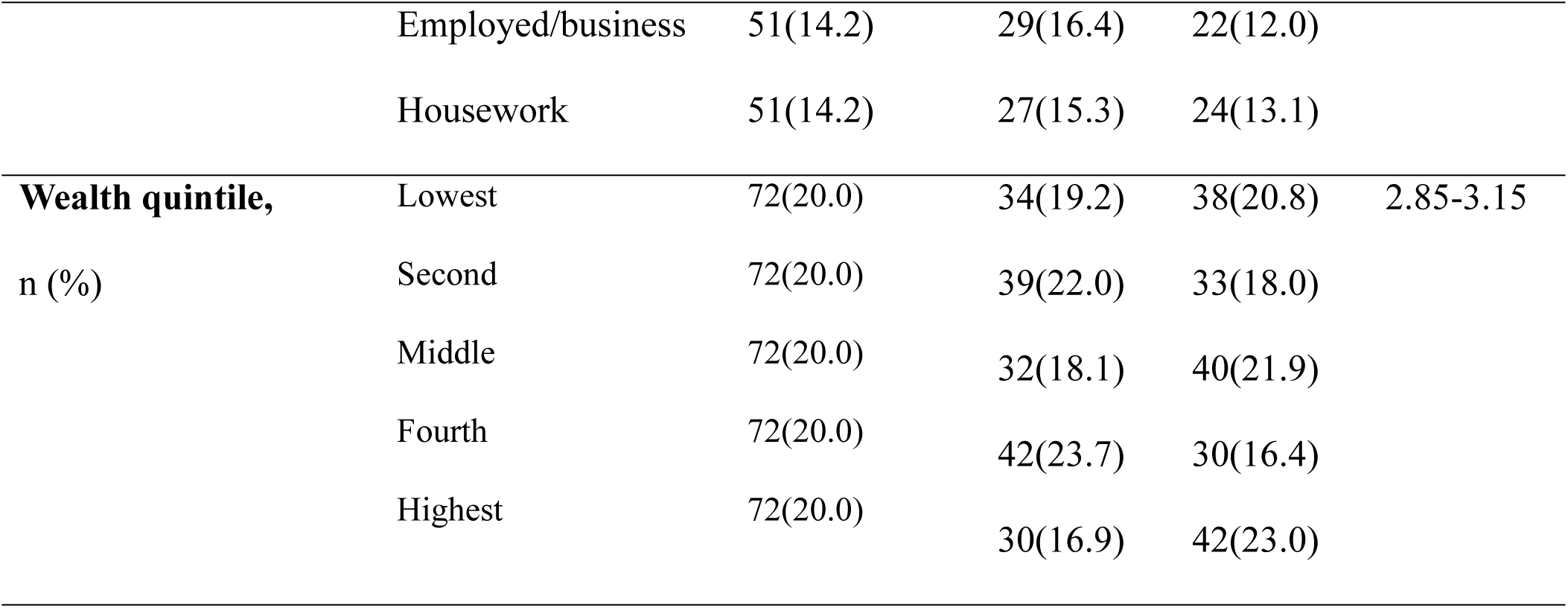
Baseline sociodemographic characteristics distributed by gender.

| Variables |  | Total(n=360) | Men<br>(n=177) | Women<br>(n=183) | 95% CI<br>(in total) |
|---|---|---|---|---|---|
| **Age class,** n (%) |  |  |  |  |  |
|  | 60-70 | 200 (55.6) | 93(52.5) | 107(58.5) | 1.53-1.68 |
|  | 71-80 | 101(28.1) | 56(31.6) | 45(24.6) |  |
|  | >80 | 59(16.4) | 28(15.8) | 31(16.9) |  |
| **Ethnicity,** n (%) |  |  |  |  |  |
|  | Newar | 243(67.5) | 51(28.8) | 46(25.1) | 1.74-1.86 |
|  | Brhamin/Chhetri | 97(26.9) | 114(64.4) | 129(70.5) |  |
|  | Janjati (other) | 14(3.9) | 8(4.5) | 6(3.3) |  |
|  | Dalit | 6(1.7) | 4(2.3) | 2(1.1) |  |
| **Marital status,** n (%) |  |  |  |  |  |
|  | Married | 212(58.9) | 141(79.7) | 71(38.8) | 1.36-1.46 |
|  | Widowed | 148(41.1) | 36(20.3) | 112(61.2) |  |
| **Education,** n (%) |  |  |  |  |  |
|  | No education | 244(67.8) | 75(42.4) | 169(92.3) | 1.50- 1.71 |
|  | Literate(informally) | 52(14.4) | 41(23.2) | 11(6.0) |  |
|  | Primary school | 25(6.9) | 23(13.0) | 2(1.1) |  |
|  | Secondary and above | 39(10.8) | 38(21.5) | 1(0.5) |  |
| **Main occupation,** n (%) |  |  |  |  |  |
|  | Agriculture | 258(71.7) | 121(68.4) | 137(74.9) | 1.34-1.50 |
|  | Employed/business | 51(14.2) | 29(16.4) | 22(12.0) |  |
|  | Housework | 51(14.2) | 27(15.3) | 24(13.1) |  |
| **Wealth quintile,** n (%) | Lowest | 72(20.0) | 34(19.2) | 38(20.8) | 2.85-3.15 |
|  | Second | 72(20.0) | 39(22.0) | 33(18.0) |  |
|  | Middle | 72(20.0) | 32(18.1) | 40(21.9) |  |
|  | Fourth | 72(20.0) | 42(23.7) | 30(16.4) |  |
|  | Highest | 72(20.0) | 30(16.9) | 42(23.0) |  |

Table 2 shows the functional disability status of the study population. About 65.7% of the elderly declared being able to perform the basic activities of daily living. Compared to women, men reported a lower level of functional disability in ADL and IADL. In IADL, men were more independent (62.7%) than women (42.1%). Overall, individuals dependent for ≥ 2 ADLs were 27.5%, and 42.8% were dependent for ≥ 2 IADLs. Severe disability was more prevalent in women as compared to men; ADL dependency for ≥ 2 activities was 34.4% and IADL dependency for ≥ 2 activities was 50.3%.

**Table 2.** Functional disability of the study sample distributed by gender.

| **Variables** | **Total**<br>**(N=360)** | **Males**<br>**(N=117)** | **Females**<br>**(N=183)** | **Test**<br>**statistics** | **P** |
|---|---|---|---|---|---|
| **ADL** |  |  |  |  |  |
| Independent | 235(65.3) | 132(74.6) | 103(56.3) | 13.30 | 0.001 |
| Dependent 1 ADL | 26(7.2) | 9(5.1) | 17(9.3) |  |  |
| Dependent for ≥ 2ADL | 99(27.5) | 36(20.3) | 63(34.4) |  |  |
| **IADL** |  |  |  |  |  |
| Independent | 188(52.2) | 111(62.7) | 77(42.1) | 17.45 | < 0.001 |
| Dependent for 1 IADL | 18(5.0) | 4(2.3) | 14(7.7) |  |  |
| Dependent for ≥ 2 IADL | 154(42.8) | 62(35.0) | 92(50.3) |  |  |

Table 3 shows the prevalence of malnutrition in the study population. Approximately 13.1% of the study population was malnourished (MNA score < 17), 45% were at risk of malnutrition (MNA score 17-23.5) and 41.9% were having normal nutrition (MNA score > 24). There was no significant difference between men and women in nutritional status.

**Table 3:** Nutritional status of the study sample distributed by gender.

| **Variables** | **Total**<br>**(n=360)** | **Men**<br>**n=177** | **Women**<br>**n=183** | **Test**<br>**statistics** | **P** |
|---|---|---|---|---|---|
| **MNA** |  |  |  |  |  |
| Malnutrition (<17) | 47(13.1) | 26(14.7) | 21(11.5) | 2.75 | 0.251 |
| At risk of MN (17-23.5) | 162(45.0) | 72(40.7) | 90(49.2) |  |  |
| Normal (>24) | 151(41.9) | 79(44.6) | 72(39.3) |  |  |
| **BMI** |  |  |  |  |  |
| Underweight (<18.5) | 42(11.7) | 26(14.7) | 16(8.7) | 6.01 | 0.110 |
| Normal weight (18.5-24.99) | 216(60.0) | 109(61.6) | 107(58.5) |  |  |
| Over weight (25-29.99) | 84(23.3) | 36(20.3) | 48(26.2) |  |  |
| Obesity (>30) | 18(5.0) | 6(3.4) | 12(6.6) |  |  |

P values are from Chi square test.

The undernutrition rate was 11.7% in the study’s BMI analysis. More underweight was seen in men (14.7%); however, overweight and obesity were higher in women, at 26.2% and 6.6% respectively.

Table 4 shows the WHO well-being index scores of the participants of the study. In the WHO well-being index score, 32.8% had a score of < 13, reflecting a condition of poor subjective psychological well-being.

**Table 4.** Psychological well-being of the study sample distributed by gender.

| **Variables** | **Total<br>(N=360)** | **Males<br>(N=117)** | **Females<br>(N=183)** | **Test statistics** | **P** |
|---|---|---|---|---|---|
| **WHO-5-A score** |  |  |  |  |  |
| < 13 | 118(32.8) | 43(24.3) | 75(41.0) | 11.37 | 0.001 |
| ≥ 13 | 242(67.2) | 134(75.7) | 108(59.0) |  |  |

P values are from Chi square test.

According to the WHO-5 A score, almost one-third of the participants had a score <13, suggesting poor psychological well-being and poor quality of life. It was also an indication for testing for depression. Approximately 41% of women reported poor subjective well-being, which is more than men (24.3%).

## Discussion

This study explored the status of functional disability, malnutrition, and psychological well-being in the elderly population of urban areas of the Lalitpur District of Nepal.

Our study findings revealed a high prevalence of disability, malnutrition, as well as poor psychological well-being among older people in the community. The prevalence of functional disability was 34.7% in ADL score and 47.8% in IADL score. An increased disability rate was observed in women. Malnutrition prevalence was 13.1% in the study population, with 45.0% at risk of malnutrition. According to the WHO well-being index score, 32.8% had poor psychological health, more so in women, needing evaluation and management of depression.

Disability is a risk to the health and autonomy of older people. Our study found that about 35% were ADL dependent. Among those, more than 27% were severely dependent according to the ADL scale. Females had a higher severity of dependence than males. Approximately 48% of the population was IADL dependent, with severe dependency at 27.5%. Based on the data of the 2011 National Census of the Lalitpur district, the reported physical disability rate was relatively low (1.05%), although academic studies suggested higher rates in older populations. However, the census categorized disabilities into several specific types: physical disability, deafness, blindness, speech problems, mental disability, and intellectual disability. In Lalitpur, the older population made up 5.32% of the total population, many of whom fell into the “Blind/Low Vision” or “Deaf” categories [46].

The national survey, NDHS 2021, revealed 27% disability in the elderly in Nepal [4], although this survey used the Washington Group Short Set of questions on functioning to assess disability. In our study, ADL and IADL tools were used to assess physical disability.

Ghimire et al. [21] found 8.3% poor functional disability among the older population in Nepal using the ADL index. Different geography, settings, and the timing of the study may have influenced the difference in this finding. However, more studies are needed to understand the prevalence and various dimensions of functional disability among older adults in Nepal.

In neighboring countries, studies have found different prevalence rates of functional disability among older individuals. Han et al. [47] assessed IADL dependency among the Chinese elderly, and IADL dependency was 38% in that study. Functionally dependent elderly have worse nutrition and health status than functionally independent older adults. In a study carried out to investigate the impact of nutritional status on physical function among elderly people in Bangladesh, Ferdous et al. [48] assessed nutritional status by MNA and disability by ADL score, performance tests, mobility, and handgrip strength. This study showed that 8% of the study population had limitations in ADL.

This result is lower compared to our study results. Physical disability caused by numerous disorders is a prevalent health problem of the elderly which can be improved by various measures. The health of the elderly population was seen to improve, and disability was reduced in the US population, as a result of improved public health measures such as improved medical treatments, use and availability of assistive devices, and behavioral changes (stopping smoking, reduced fat intake) in the elderly [49].

Our study found a 13.1% malnutrition prevalence among older people. Malnutrition is associated with an increased risk of cardiac and respiratory problems, infections, deep vein thrombosis, pressure ulcers, perioperative mortality, and multi-organ failure [50].

Malnutrition in older adults is multifactorial. Sensory decline, such as reduced taste and smell, makes food less palatable. Inflammation from chronic diseases (cancer, heart failure) increases the body’s nutrient requirements while simultaneously suppressing appetite [51].

Oral health conditions, such as poor dentition, ill-fitting dentures, or dry mouth, make chewing and swallowing difficult [52]. Cognitive decline can lead to forgetting to eat or the inability to recognize food, leading to malnutrition. Additionally, social isolation is one of the strongest predictors of nutritional risk [23].

Many medications cause side effects like nausea, altered taste, or reduced nutrient absorption, early satiety, anorexia, dry mouth, and taste impairment [51]. Overly strict diets for diabetes or hypertension can sometimes lead to an accidental calorie deficit.

Decreased functional ability is another cause of malnutrition in older adults because it may limit their gross body movement and self-care [53]. Malnourished patients have extended lengths of stay when hospitalized and have more complications compared to patients of normal weight [51]. As a result, an increased burden of healthcare costs is associated with undernutrition and malnutrition. In an aging society, the importance of early nutritional assessment in older adults is noteworthy because of its profound therapeutic, prognostic, and health maintenance implications [54].

Health-protective effects of psychological well-being are evidenced in various research studies. This subjective well-being reduces the risk of disease and promotes longer life [25]. In our study, we found 32.8% of older individuals had poor psychological well-being. More men (75.7%) had a good quality of life and subjective well-being than women (59.0%). Women were significantly more likely to have mood symptoms and a poor quality of life than men (p = 0.001).

The essential features of well-being are: purpose in life, autonomy, personal growth, environmental mastery, positive relationships, and self-acceptance. These variables provide some indicators based on feelings of goodness, joy, positivity, or satisfaction with life [25].

The WHO-5 Well-Being Index focuses on positive moods, such as feeling calm, active, and rested in the last 2 weeks (short-term state). It measures the affective and hedonic dimensions of an individual’s well-being. This tool assesses the hedonic balance, which is a relation between positive and negative emotions. Furthermore, this evaluates the duration and intensity of emotions [55].

In our study, nearly one-third of the study participants reported poor psychological well-being. Ghimire et al. [21] reported 21% poor life satisfaction among the elderly using the Satisfaction with Life Scale (SWLS) in Nepal. The 5-item SWLS (5-35 range) measures a person’s cognitive judgment of their life satisfaction, rather than emotional feelings, while the 5-item WHO-5 assesses psychological well-being in an individual.

The higher prevalence of widowhood in our study might be one of the reasons for increased poor psychological well-being, especially in women. In our study, 61.2% of women were widowed compared to 20.3% of men. Anxiety, depression, and stress are common among Nepalese widows; also, older age has been associated with a poor mental health state [56]. These factors, as well, may have influenced the findings in our study.

In addition, increased age and a lack of formal education were associated with depression in Nepal [57]. In our study population, the illiteracy rate among older individuals was 67.8%, and 92.3% of women were illiterate, which is comparatively higher than the rate for men.

Challenges in life, such as frailty, disability, loss of a spouse, social inequalities, or unexpected traumas, can result in a low quality of life and poor psychological well-being [58]. In such circumstances, obviously, well-being is compromised.

Interventions have improved the well-being status of individuals, reduced mood symptoms, and have impacted them to live longer with greater resilience. Resilience is the capacity to maintain well-being in the face of adversity [25].

### Strengths and limitations

This study reported the functional disability status, prevalence of malnutrition, and psychological well-being with gender differences in older adults in the urban population of the Lalitpur district of Nepal. Some characteristics were self-reported, so there may be a likelihood of recall bias. There is a time gap between the data collection and the publication of the findings; due to various circumstances, the findings reflect the status of older people at the time of the survey. Knowledge of the risk factors for disability, malnutrition, and psychological well-being would have provided more comprehensive results in the study among older adults.

### Future policy and practice prospects

Poorly equipped healthcare facilities and the lack of a competent workforce trained in geriatric care make it challenging to provide quality care to older adults in Nepal. It is now mandatory to dedicate geriatric units in hospitals in Nepal, yet compliance is poor [59]. The geriatric workforce is almost non-existent, and medical education in Nepal does not offer geriatric specialization, further exacerbating the workforce shortages [60]. Day centers and old-age homes are scarce, geographically limited, and expensive. The care provided is not regulated or monitored [59].

Additionally, the absence of rehabilitation, palliative care, assisted living, and skilled nursing facilities points to the urgent need to build such facilities, especially given the rapid growth of the older adult population with increased comorbidities [60].

This study indicates the urgency of public health measures for functional disability prevention and management among older people in the community. Healthcare for older adults must be focused on the prevention of functional disability, malnutrition, and poor mental health.

Effective rehabilitation with a team of physiotherapists can increase mobility and functional outcomes. Occupational therapists can improve ADLs, audiologists can take care of hearing problems, nutritionists can manage undernutrition, and healthcare facilities can manage morbidity such as diseases and mental health issues.

Geriatric healthcare needs assessments, such as comprehensive geriatric assessment (CGA), should be in place and practiced. The assessment of the core elements of CGA, such as disability, social support, disease severity, comorbidities assessment and management, nutritional assessment, mental health care, and cognition, should be performed in a timely manner.Rehabilitation with a multi-team approach, community mental health services, social networks, and education and management of undernutrition are essential for elderly well-being.

Public health policies aimed at managing functional disability, mental health, and malnutrition are crucial for elderly well-being. Furthermore, older people need more assistance from others and from the health care system as well. People having these health conditions may improve, and ultimately this will decrease the burden of ill health on both the individual and society.

### Recommendations

There is evidence that effective rehabilitation improves functional status and decreases mortality. Multidisciplinary occupational therapy is an effective measure to take care of the health of older people. This team approach should be planned and practiced in healthcare facilities.

Implementation of CGA in healthcare delivery facilities is crucial, and the intervention measures, as mentioned previously, should be utilized widely.

Interventions for poor psychological well-being and malnutrition should be accessible to those who are vulnerable in the community, especially focusing on women. The healthcare delivery system should be elderly-friendly with health service providers trained in the health of older adults.

### Conclusions

This study assessed the main components of geriatric health: functional disability, psychological well-being, and malnutrition. The results of the study revealed an increased prevalence of functional disability, poor psychological well-being, and malnutrition in the older population.

The functional disability rate was high among older people. Nearly one-third of the participants were dependent on one or more activities of daily living. About one-third of the study population was in a state of poor psychological well-being. And about half of the population had poor nutritional status. The prevalence of functional disability and poor psychological well-being was high in women.

This study highlights the need to address the health of older people, focusing on geriatric assessments to identify at-risk older individuals. Plans and programs should be developed for prevention and targeted interventions for this vulnerable group.

## Authors’ contributions

Conceptualization: Eebaraj Simkhada, Aerusha Simkhada

Formal analysis: Eebaraj Simkhada

Investigation: Eebaraj Simkhada, Aerusha Simkhada

Supervision: Eebaraj Simkhada.

Writing – original draft: Eebaraj Simkhada.

Writing – review & editing: Eebaraj Simkhada, Aerusha Simkhada

## Data availability

The dataset supporting the conclusions of this article is included within the article. The data are available upon request from the corresponding author.

## Ethics approval and consent to participate

This study was approved by the Institutional Review Committee, Institute of Medicine, Maharajgunj Medical Campus, Tribhuvan University, Nepal (Ref. no. 75(6-11-E)071/072). This study followed the ethical standards laid down in the 1964 Declaration of Helsinki. All participants were informed of the study’s subject and method. Written consent was obtained from all participants, and thumb impressions were obtained from illiterate participants.

